# An Exploration of Impact of COVID 19 on mental health -Analysis of tweets using Natural Language Processing techniques

**DOI:** 10.1101/2020.07.30.20165571

**Authors:** Sohini Sengupta, Sareeta Mugde, Garima Sharma

## Abstract

Twitter is one of the world’s biggest social media platforms for hosting abundant number of user-generated posts. It is considered as a gold mine of data. Majority of the tweets are public and thereby pullable unlike other social media platforms. In this paper we are analyzing the topics related to mental health that are recently (June, 2020) been discussed on Twitter. Also amidst the on-going pandemic, we are going to find out if covid-19 emerges as one of the factors impacting mental health. Further we are going to do an overall sentiment analysis to better understand the emotions of users.

**Executive Summery:** Novel Corona virus’s spread and its impact on various aspects of national and individual’s well-being has been at the center of lot of discussions across micro-blogging sites and various social media platforms ever since it commenced in December 2019. Users are voicing their opinions on several topics related to covid-19. Social distancing as prescribed by Government and Local Administration We all are aware that the Novel Corona virus has significantly affected our physical health; however the current social distancing norms are taking a toll on the psychological well-being of individuals. The research paper presents a two-phased analysis of most recent 2000 tweets related to mental health pulled out twice over a span of one month on 28 June 2020 and 28 July2020 respectively, thereby analyzing 4000 tweets in total. The second phase analysis was conducted exactly after a gap of one month to validate the results generated by the first analysis. The intention is to analyze to what extent people have discussed about mental health in the past few months based on the information disseminated on Twitter. Data was extracted using Twitter’s search application programming interface (API) and Python’s tweepy library. A predefined keyword like ‘mental health’ was given to find out if Covid-19 emerges as a reason for the same. Several natural language processing (NLP) techniques like tokenization, removing URL and stop words, stemming and lemmatization were used to pre-process the text data and make it ready for analysis. These collected tweets were analyzed using word frequencies of single and double words (unigram, bigram). A very unique feature of this analysis includes a network diagram that shows interconnections between the set of most common words used in to its and the connections (if any) are represented through links. Topic modeling technique in NLP visualizes the top concerns of tweeters through a word cloud. At present we have many methods to do topic modeling. In this paper we are using the Latent Dirichlet Allocation (LDA) method which is a probabilistic approach of modeling given by Prof David H.B in 2003. This model deals with distribution of topics to tweets and allocation of those topics to documents and words to topics. Finally a sentiment analysis is done using text mining techniques to analyze the sentiment of the tweets in the form of positive, negative and neutral.

## Introduction

In Twitter we can extract certain metadata like source of tweets, location of tweets and also the individuals involved. Retweets and replies can also be studied. This post closely reflects user’s concerns, reactions and emotions. Also feature allows users to post in different languages and in different forms like links, images, videos and texts. This paper includes analysis of only text data in English language. With every passing day, mental health is becoming a more common issue. WHO (World Health Organization) supports this by stating that one out of every four individuals is bound to face mental health disorders at some point in their life. Apart from the risk of the virus infection, the covid-19 pandemic has brought quite a number of social outcomes like financial crisis, job loss, increasing unemployment to mention a few. Sentiment Analysis of all these issues can be done by using twitter data. People involved in essential services are more likely to experience psychological stress. However to figure out the solution to this we need to understand the mental state first. People express their thoughts and opinions in several social media platforms like Facebook, Instagram, Twitter, Reddit etc. The current pandemic that forced huge number of people to work in a virtual environment has been a huge driver for surge in active social media users. (Source-Statistica.com)

### Review of Related Literature

Twitter allows collection of its data by a standard search application program interface (API). This generates tokens and then we can extract tweets on any topic. A lot of people have done Twitter data analysis considering several factors related to covid-19. Facebook and some other social media platforms also allow us to do a sentiment analysis of user’s posts. All this analysis used techniques like Natural Language Processing (NLP), text mining and thereby identify the overall common responses of people towards the pandemic. [1]Alrazaq, Alhuwail, Housen, Hamdi (2020) conducted a study using similar techniques by extracting tweets related to covid-19. They performed sentiment analysis and came out with 4 broad topics that were most discussed, namely –

a. Origin of COVID-19
b. Source of novel Coronavirus
c. Impact of Covid-19 on people and countries
d. Methods for decreasing the spread of Covid-19.

They used an unsupervised machine learning technique called Topic Modeling that is capable of forming clusters in the collection of tweets. The algorithm used was Latent Dirichlet Allocation from Python scikit learn library. [2] Ahmad and Murad (2020) intend to determine the spread of panic due to Covid-19 in Iraq. The study reveals the impact of social media panic depends on factors like education level, age, gender etc. Also their analysis concluded that individuals within the age group of 18 to 35 are more likely to be suffering from anxiety and depression. [3] Prior to the emergence of Covid-19 pandemic also researchers have used Twitter data to study impact of social media on mental health. One such study can be found in a paper ‘Enhancing the positive impact of social media on our mental health-2019’ in ‘Sage Journals’. This paper reveals how youths in the age bracket of 14 to 24 have found social media to be a reason for their possessing a good mental health and well-being. Different pages and groups on social media have given them support in their tough times. [4]There have also been fairly detailed studies on the negative impacts of social media – how fake news creates panic among individuals thereby affecting their mental health. With regards to fake news and negative impacts covid-19 related tweets are no exception. In fact in certain cases the social media panic seems to have traveled faster than the actual outbreak which may perhaps be owing to the fact that a large proportion of population does not check the authenticity of information source. A similar study conducted by Kadam and Etre (2020) is documented in their paper ‘Negative impact of social media panic during the covid-19 outbreak in India-2020’ published in ‘Journal of Travel Medicine’. [5] As far as authenticity of sources of content in social media is concerned, the role of policymakers assumes huge importance. Fake news detection and its removal there-after is done actively by back-end engineers. In fact social media platforms like Facebook, Instagram, Twitter have become each other’s competitors in the space of identification of news from non-reliable sources and the extent to which each one of them is successful and fast in doing so. Analysis of posts meant to hurt religious sentiments or offensive to a political party or an individual or hurting a whole community for that matter by creating panic accounts to mapping social behavior, public sentiment and subjective thoughts on a larger scale. Cinelli, Quattrociocchi and Galeazzi (2020) tried capturing this in their paper ‘The covid-19 social media infodemic-2020’ in the Pre-print journal ‘arXiv’. [6] Li, Awarez, Gasulla (2020) dwell on effects of Covid-19 on mental health. The study trained deep learning models which would be able to classify tweets in different emotions like joy, sadness, anger, fear etc. Their results showed most of the tweets are related to sadness and fear and the impact of covid-19 was also well explained. [7]Another ill-effect which emerged strongly in the times of covid-19 pandemic is stigmatization. Budhwani (2020) captures the impact of Stigmatization on mental health using quantitative analysis.

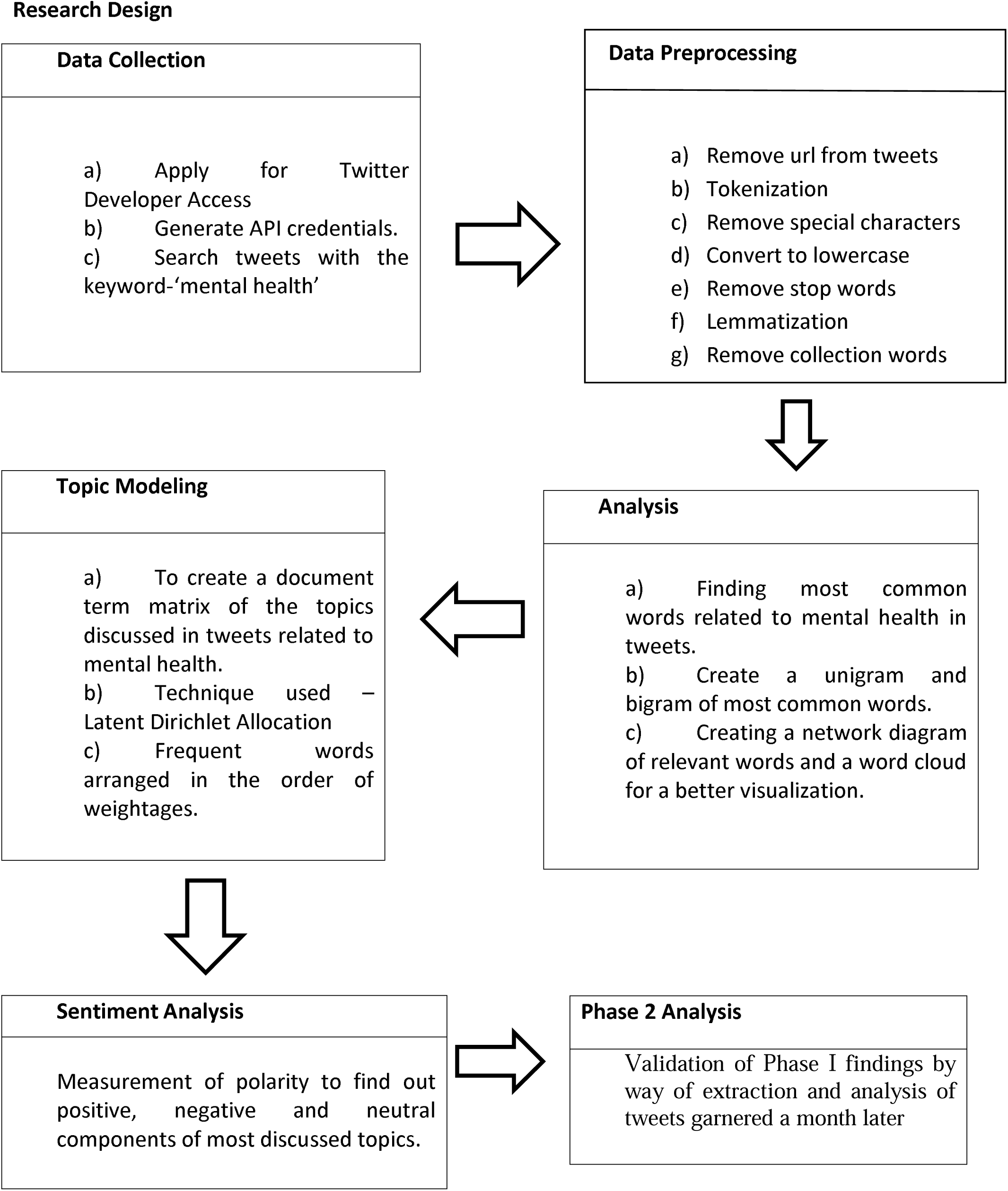

## Discussion and Analysis

In this paper we will be performing topic modeling on Twitter data in order to figure out what exactly people are tweeting regarding mental-health in these times of Covid-19 pandemic crisis. First for extraction of data, one has to have access to the Twitter API. A twitter developer account was created for generation of credentials viz. ‘Consumer key, ‘consumer secret’, ‘access token’ and ‘access token secret’. Passing those generated credentials tweepy’s OAuth Handler named as ‘auth’ in Python, we can get complete access to each and every tweet.

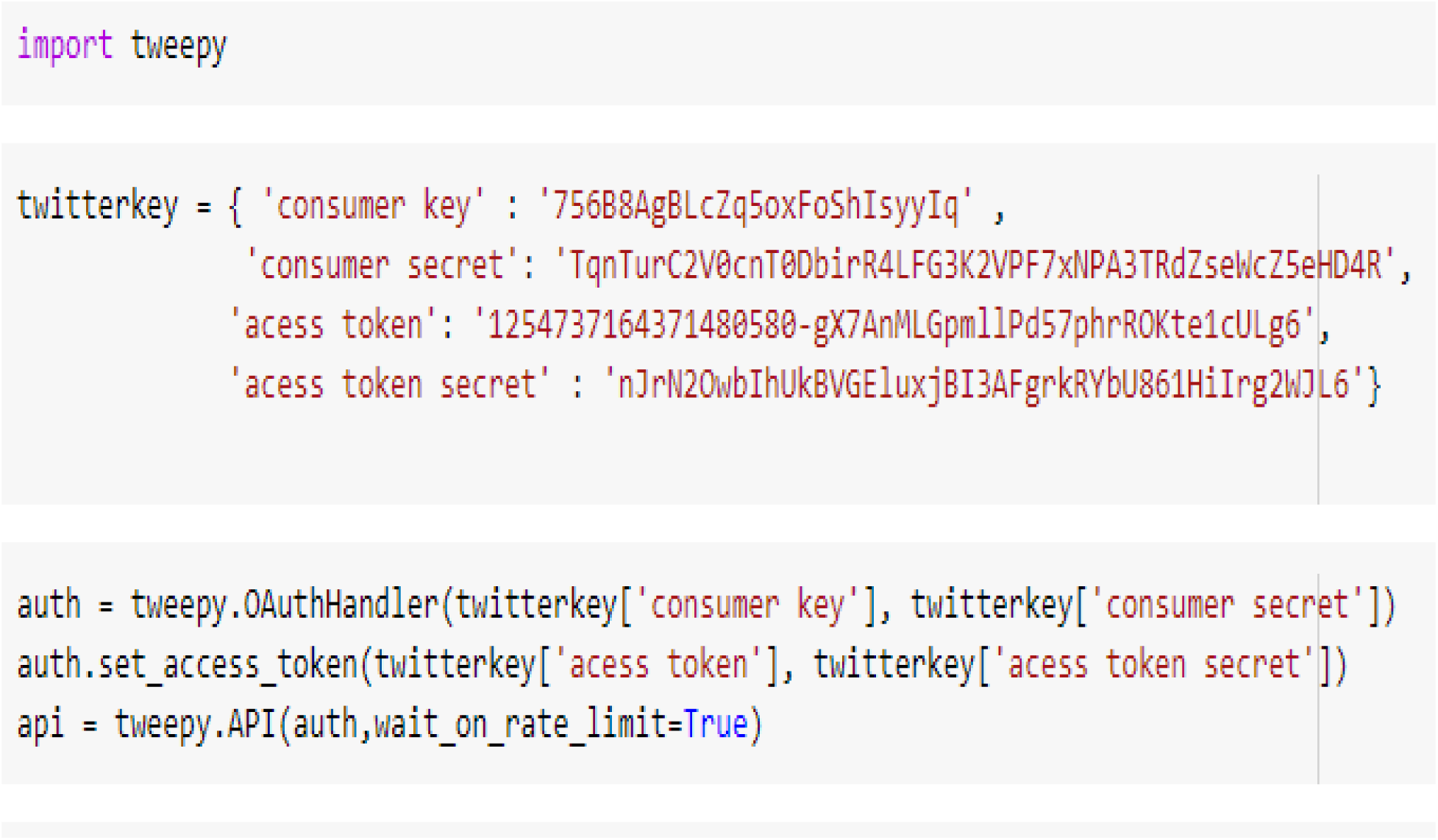

The keyword ‘mental-health’ was identified to study its related tweets. Thereafter 2000 odd most recent tweets were extracted on 28^th^ June, 2020 which then underwent certain data preprocessing techniques like removal of URL, tokenization, removing stop words, lemmatization and converting all letters into lowercase. Finally data was incorporated into a data frame to make it ready for analysis.

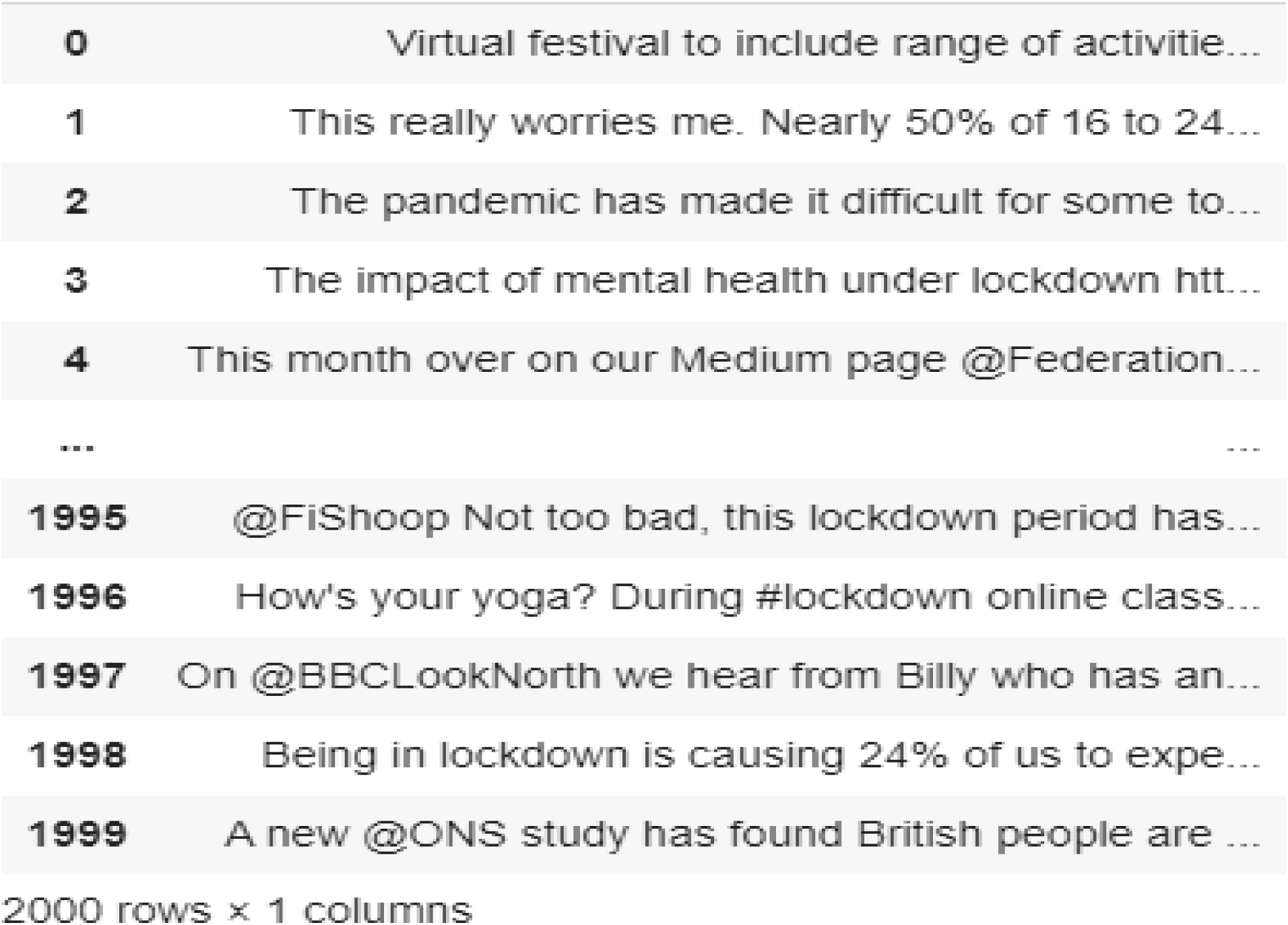

Snap Shot of Tweets Extracted In First Phase

Most common words used in twitter were found by way of both unigram and bigram analysis. Unigram takes into consideration single word. Bigram considers two consecutive words that are most used. Bigram analysis is more contextual in understanding any topic.

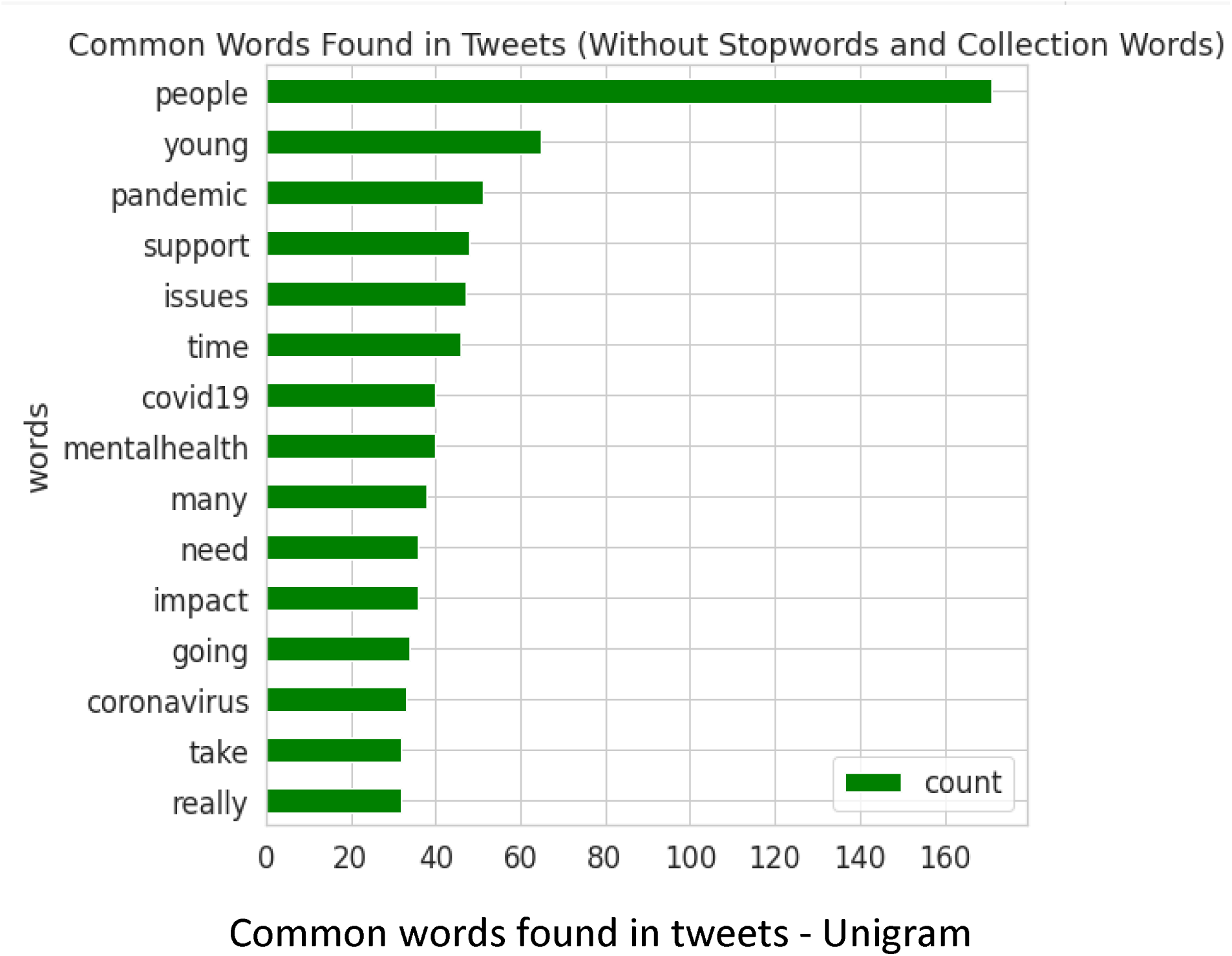

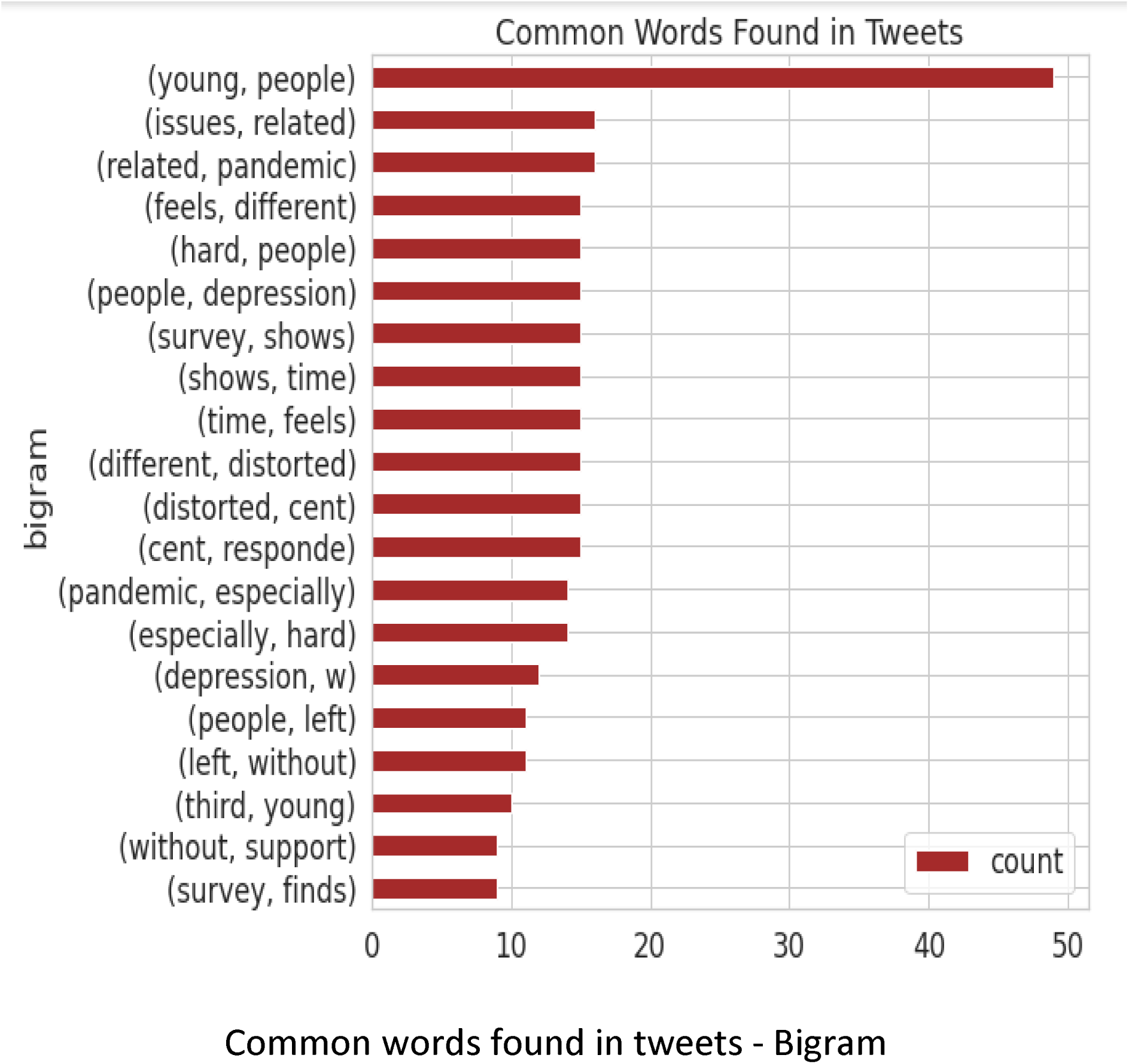

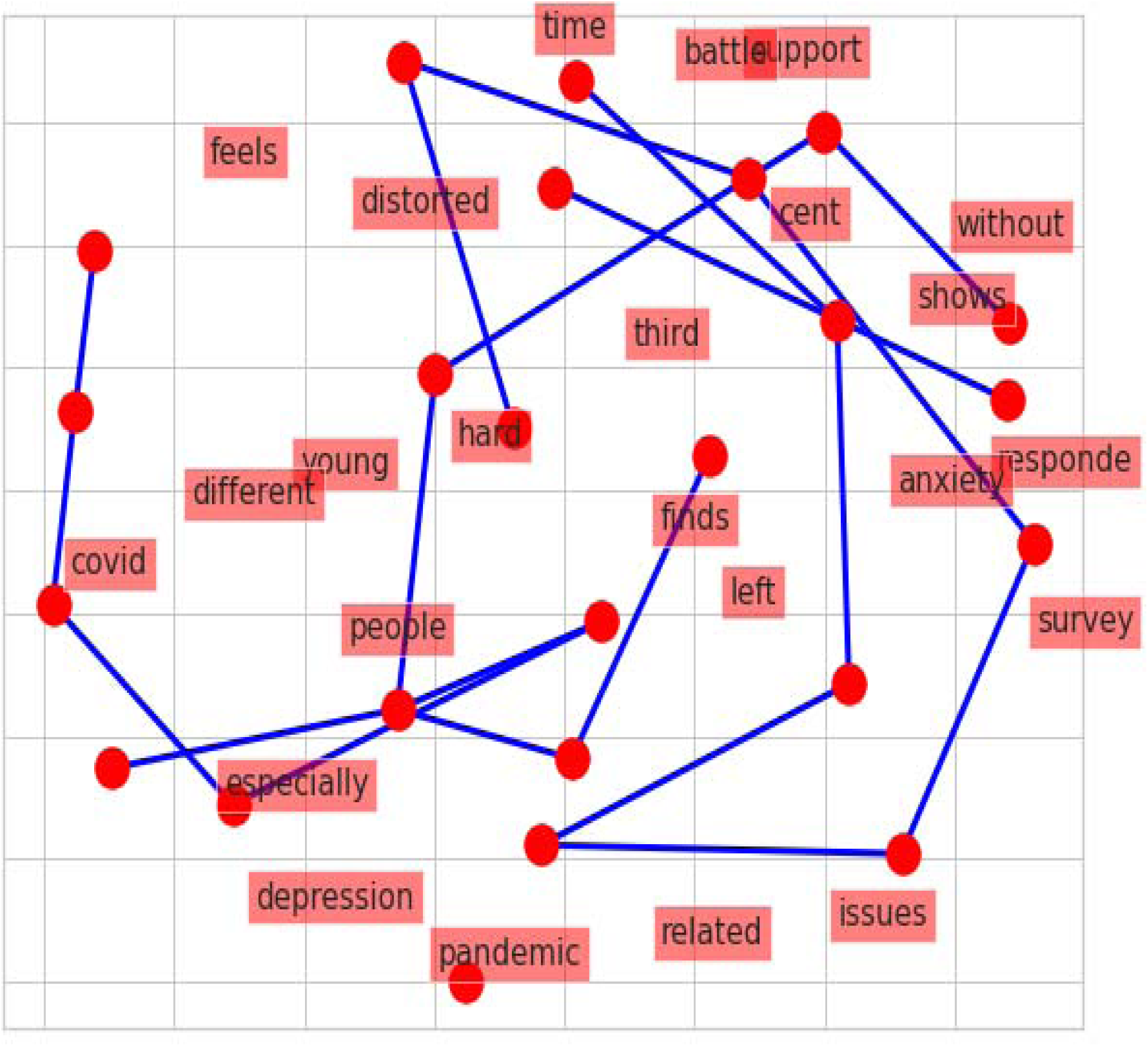

A network diagram to visualize keywords and their relationships. Each word is connected with an Arc.

Word cloud is a text mining technique that aids in construction of a story line. It consists of a large number of individual words which in isolation sometimes does not make much sense, however, a lot of words related together can draw a million-dollar story. Hence the need to identify texts so as to extract words from tweets. Thereafter a document term matrix was built on the already cleaned and preprocessed data. This enabled highlighting of the most commonly and frequently cited words in all the tweets related to mental health. It is one of the best ways to visualize relevant keywords on a chosen topic.

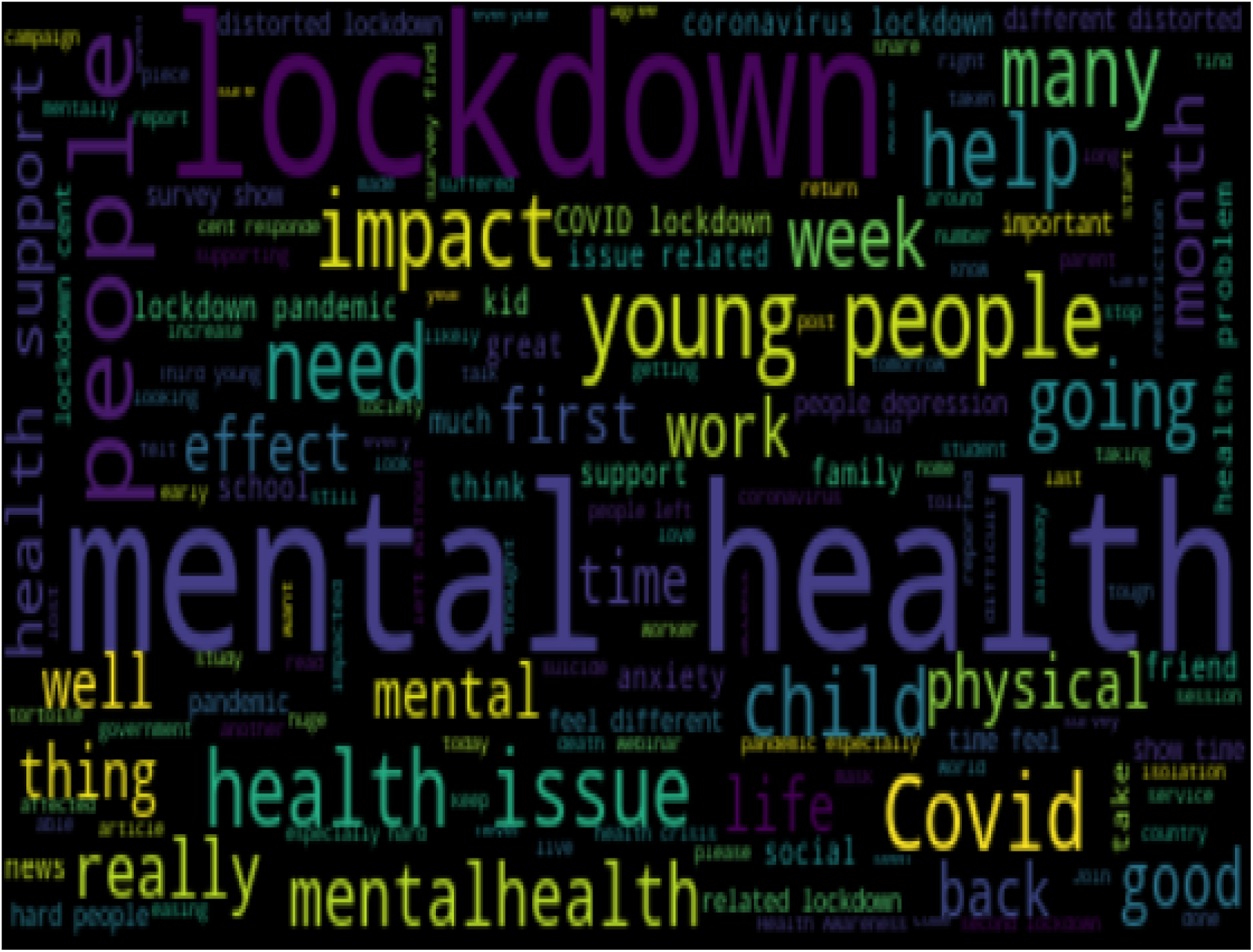

Topic modeling - It is an unsupervised NLP technique that can be used to visualize a text document by studying the topics of discussion present there. It is almost same as the clustering feature of machine learning the only difference being that here we are capturing collection of words from text data tweets instead of numerical data. Let the number of topics chosen be 10. Python randomly chooses different topics from the entire distribution and randomly assigns weights to them. After this LDA model uses Gibbs sampling method to do iterations in giving weightages. What this algorithm basically does is gives highest weightage to those words that have the maximum conditional probability.

|  | Topic 0 words | Topic 0 weights | Topic 1 words | Topic 1 weights | Topic 2 words | Topic 2 weights | Topic 3 words | Topic 3 weights | Topic 4 words | Topic 4 weights |
| --- | --- | --- | --- | --- | --- | --- | --- | --- | --- | --- |
| 0 | coronavirus | 1.2 | take | 0.2 | mentalhealth | 1.2 | time | 1.2 | impact | 1.2 |
| 1 | going | 1.2 | pandemic | 0.2 | many | 1.2 | need | 1.2 | covid19 | 1.2 |
| 2 | young | 1.2 | support | 0.2 | support | 1.2 | really | 1.2 | people | 1.2 |
| 3 | take | 0.2 | many | 0.2 | pandemic | 1.2 | issues | 1.2 | take | 0.2 |
| 4 | pandemic | 0.2 | mentalhealth | 0.2 | take | 1.2 | pandemic | 0.2 | pandemic | 0.2 |

The keywords in each topic seem to be repeated with the number of topics is equals to 10. A trial and error method is used to find the suitable number of topics-it is 2 in our case.

|  | Topic 0 words | Topic 0 weights | Topic 1 words | Topic 1 weights |
| --- | --- | --- | --- | --- |
| 0 | coronavirus | 1.5 | take | 1.5 |
| 1 | issues | 1.5 | impact | 1.5 |
| 2 | going | 1.5 | pandemic | 1.5 |
| 3 | support | 1.5 | really | 1.5 |
| 4 | covid19 | 1.5 | mentalhealth | 1.5 |
| 5 | young | 1.5 | people | 1.5 |
| 6 | time | 1.5 | many | 1.5 |
| 7 | need | 1.5 | need | 0.5 |
| 8 | many | 0.5 | time | 0.5 |
| 9 | people | 0.5 | young | 0.5 |

Sentiment analysis-This is to determine if a piece of tweet/text data is positive, negative or neutral. Textblob library of python was used and NLTK corpora was installed for sentiment analysis. Polarity method of textblob was used to get the polarity of tweets between −1 to + 1. If polarity is greater than 0, tweets are positive; if it is less than zero then tweets carry a negative sentiment. A polarity value equal to zero represents neutral sentiments. Compiling this for all tweets, the result can be summarized as-

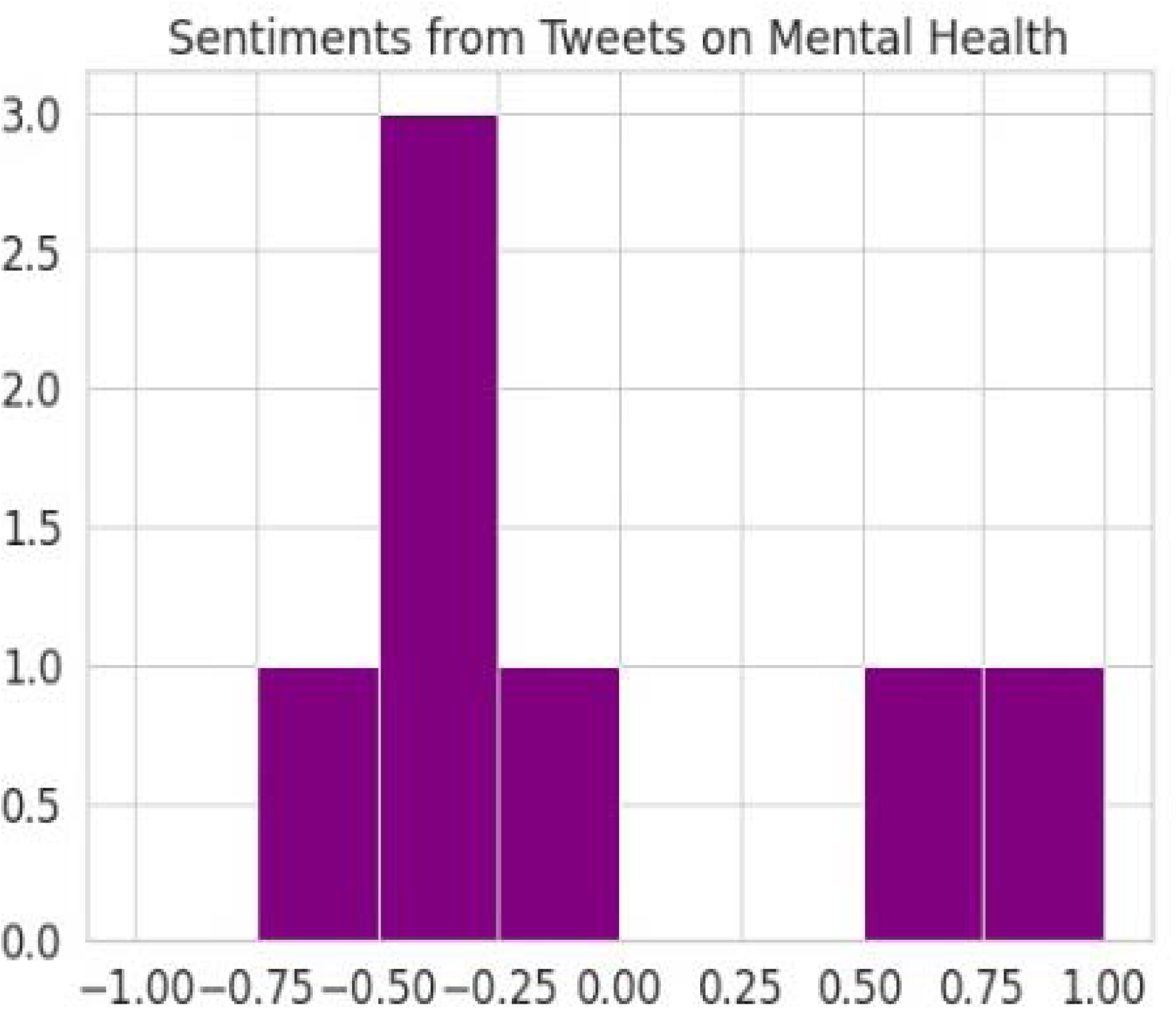

A month later on 28^th^ July, 2020 again a set of 2000 tweets were extracted for conducting the second set of analysis so as to validate the results generated by the first analysis. Some results to note are –

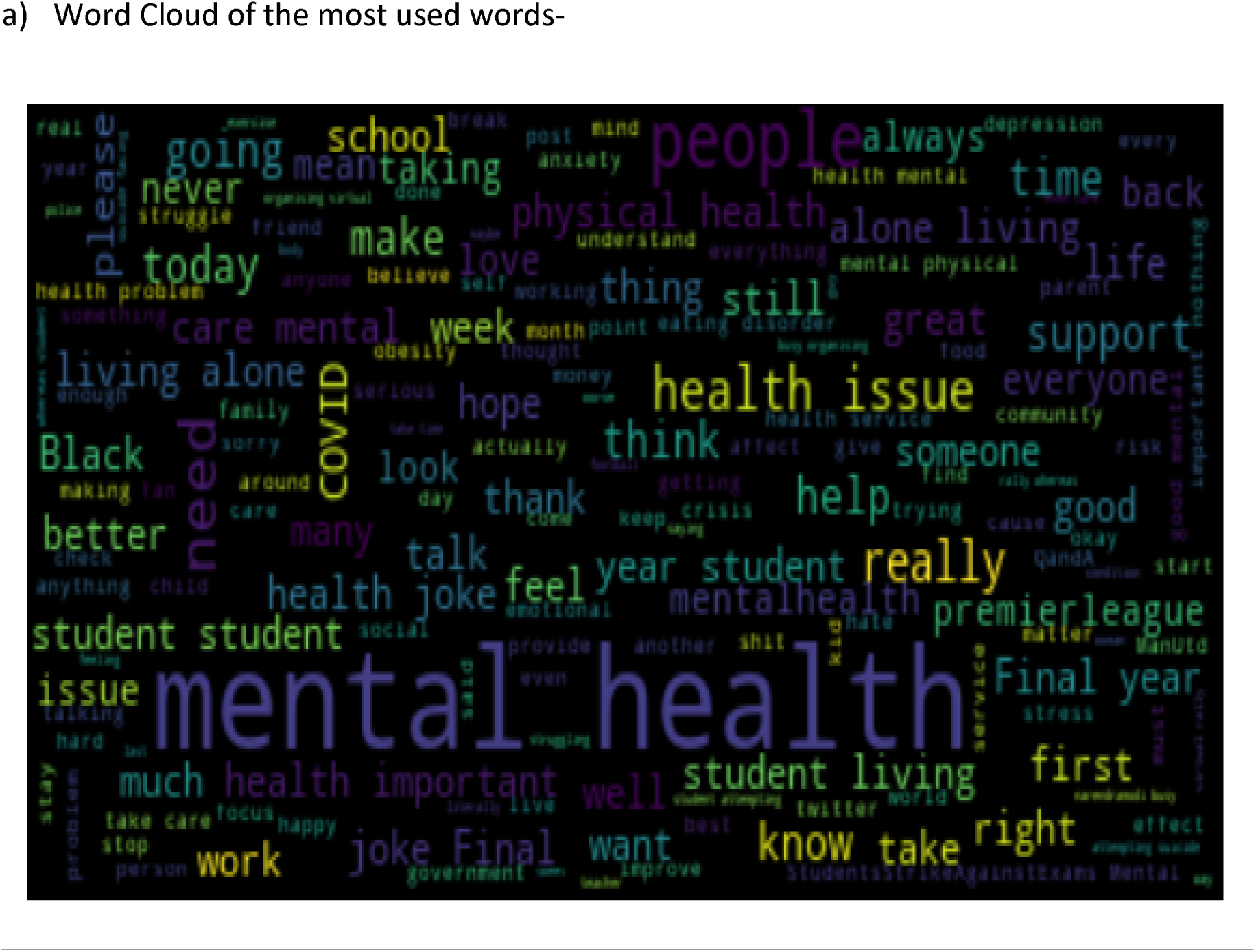

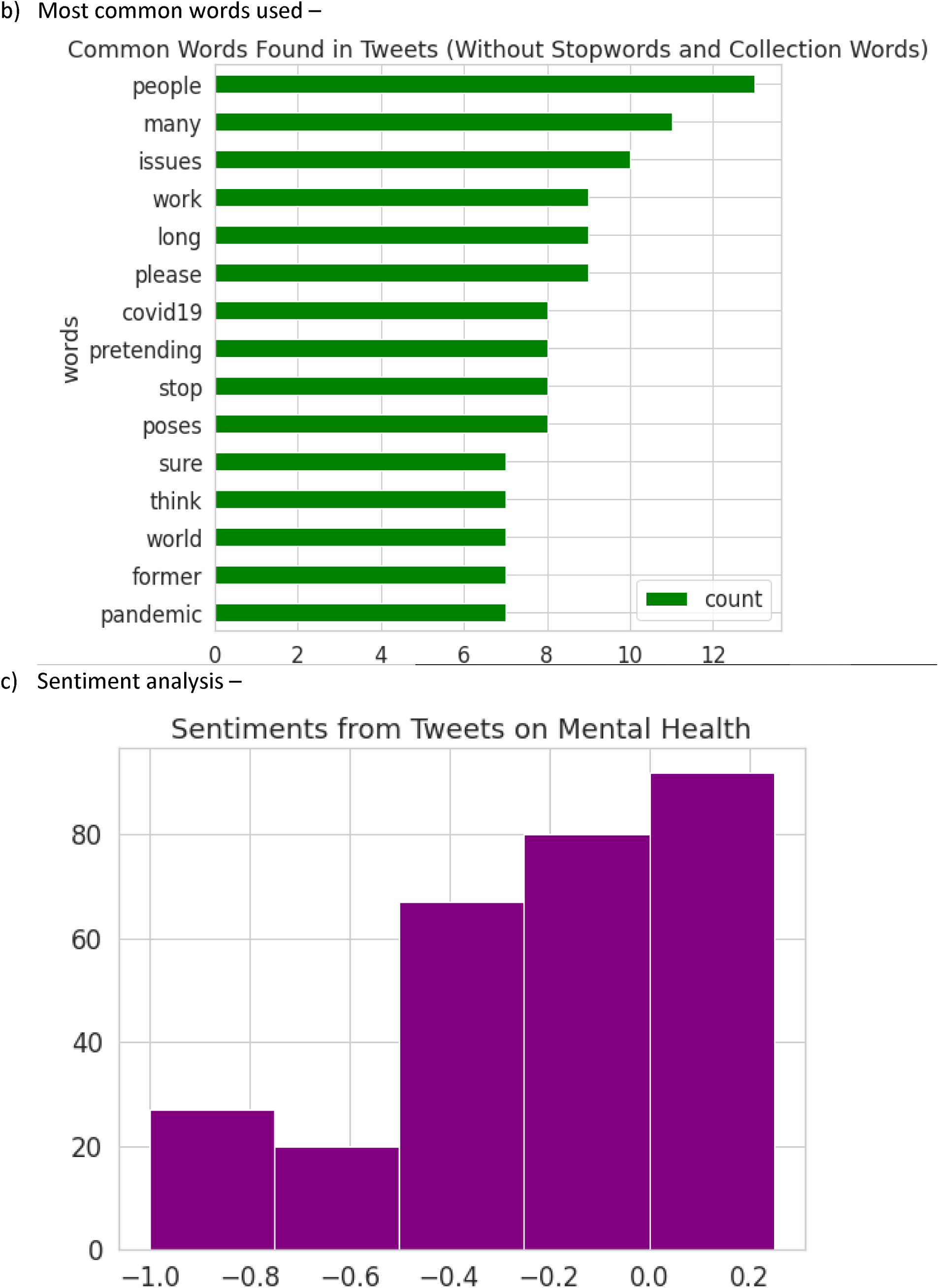

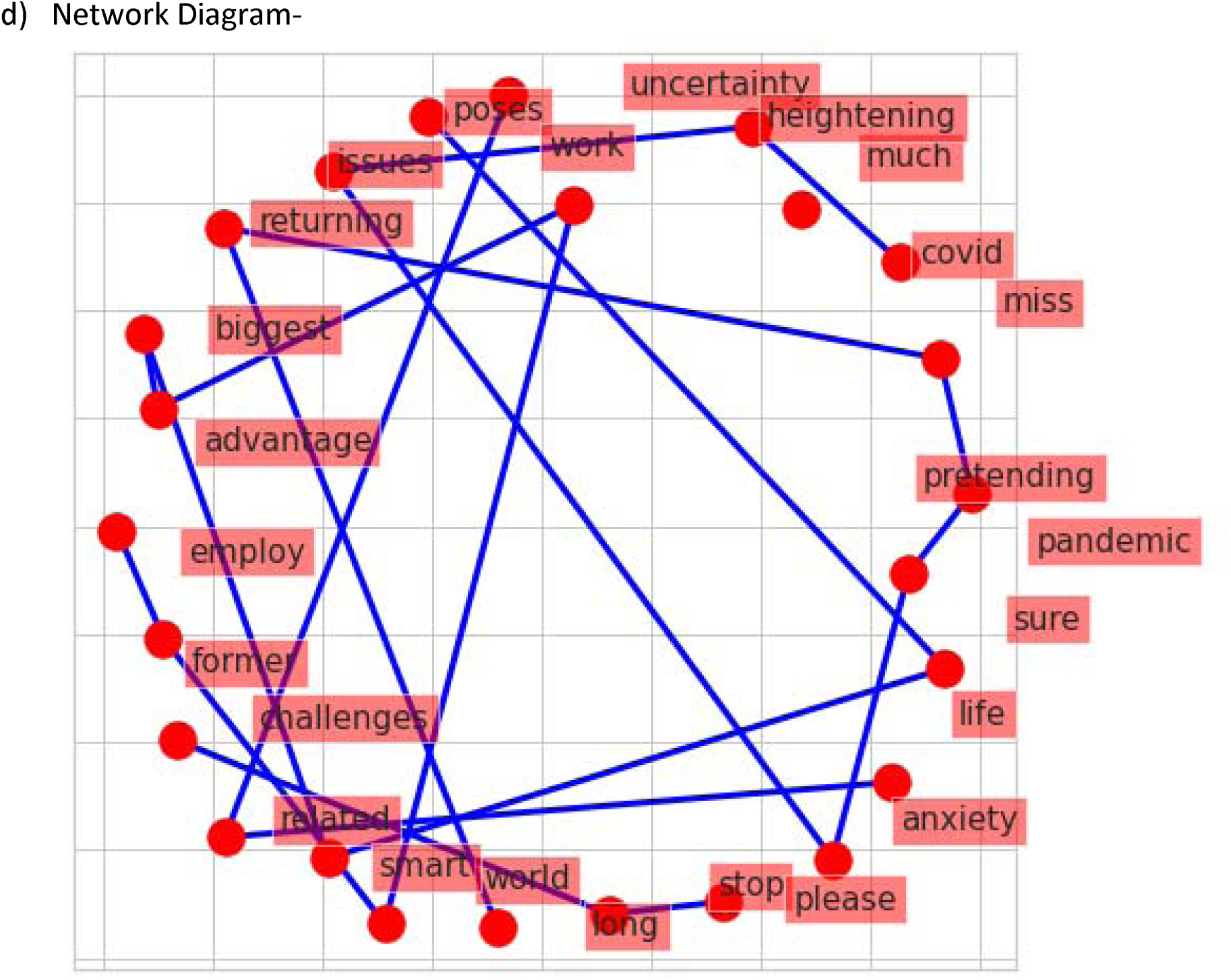

## Results

### PHASE I Analysis of tweets extracted on 28/06/2020

The unigram as well as bigram analysis of the most frequently used words shows that Covid-19 or coronavirus is one of the major current reason behind people facing mental health issues. We had used the keyword “mental health” while extracting tweets. The result in both word cloud and unigram shows that the current pandemic has had a huge impact on mental health of people worldwide. Lockdown is another frequently used word. This tells us that a lot of people staying back at their homes due to lockdown have significantly voiced their opinions regarding mental health.

Another astounding fact revealed by this analysis is that the most used couple of word basis bigram is “young people”. We also witnessed the presence of “survey” as a frequently used word. This leads to the conclusion that according to different surveys, young people are more likely to face mental health issues, depression, anxiety and they need the maximum support amidst the pandemic. A similar story can be drawn from the network diagram. The emergence of Corona virus pandemic witnessed an analogous rise in depression and anxiety. Topic modeling using the LDA method confirms the emergence of below two topics as also concluded from the two-phase analysis. To summarize-

a. The impact of covid-19 on mental health, depression and anxiety
b. A special mention of young people suffering from mental health issues.

An overall sentiment analysis reveals that the majority of the tweets have a polarity below one that is there found to have negative sentiments.

### Phase II Validation of tweets extracted on 28/07/2020

The world cloud raises similar concerns related to mental health as were evidenced in the first phase. The word “anxiety”, “Covid”, “pandemic” is coming on the network diagram of both the tests. Also the network diagram highlights the distress and uncertainty issues regarding employment among young people. Two major things that can be pointed out are – the impact of Covid-19 on mental health and also the significant impact of mental health on young people including students and working age group. Sentiment analysis again proves that the overall impact has been negative.

The fact that both the tests lead us to similar conclusions only validates our findings.

## Conclusion

Text data is usually unstructured. In our daily life we come across numerous text data in newspapers, research papers, social media etc. Like numeric data we can also take a crucial decisions based on text data. There is a hidden theme structure in every text data. In text mining analysis we focus on this. The significance of the technique deployed, namely Latent Dirichlet Allocation (LDA) can be gauged from its name itself. Latent means hidden. Here we are talking about undercurrent themes or topics which are concealed. Dirichlet is based on the concept of “distribution of distributions”. Here we are looking at distribution of topics in tweets and also distribution of most commonly used keywords in these topics. There are several techniques available for topic modeling, which if used may present similar or different results. This may perhaps trigger researches based on other techniques as well. Any research enthusiast may take up study on similar lines to that extent. Also Twitter allows users to post in different languages. The paper restricts itself to analysis of tweets in English language only. The study can be taken forward considering other languages as well. This will help us with better understanding of sentiments. This study will also be helpful to researchers who wish to find a connect between the effect of Covid-19 on mental health with other crisis situations like job loss and unemployment.

## Data Availability

All the references are given in the manuscript itself.

